# Impact of previous COVID-19 on immune response after a single dose of BNT162b2 SARS-CoV-2 vaccine

**DOI:** 10.1101/2021.03.08.21253065

**Authors:** María Velasco, Ma Isabel Galán, Ma Luisa Casas, Elia Pérez-Fernández, Diana Martínez-Ponce, Beatriz González-Piñeiro, Virgilio Castilla, Carlos Guijarro, Working Group Alcorcón COVID-19

**Author notes:** **Corresponding author**, María Velasco, Infectious Diseases. Research Unit. Hospital Universitario Fundación Alcorcón. C/Budapest nº1, 28922 Alcorcón.Madrid. Spain. **Alternate corresponding author**. Carlos Guijarro, Professor of Medicine. Rey Juan Carlos University. Internal Medicine Unit. Research, Innovation and Education Coordinator. Hospital Universitario Fundación Alcorcón. C/Budapest n°1, 28922 Alcorcón.Madrid. Spain.

## Abstract

Immune response after a single-dose of BNT162b2 vaccine is markedly increased in subjects with previous SARS-CoV-2 infection, reaching similar IgG titers to those elicited by the full two-doses in naïve cases, and increased modestly after the second dose. These data may inform the priority of the boosting dose.

## Introduction

Vaccination against SARS-CoV-2 is proposed by the WHO as the main option, at the present time, to control the COVID-19 pandemic. However, limited vaccine supply reduces the ability to reach a substantial proportion of individuals and delay the expectations of herd immunity[1]. The standard vaccination protocol for mRNA vaccines includes a 2 dose vaccine administration 3-4 weeks apart[2]. In this setting, a strategy to use single-dose of mRNA vaccines for some individuals could accelerate the achievement of this goal. Data on immune responses to single-dose vaccination with BNT162b2 are limited to few observational studies with small number of patients [3–6]. Moreover, impact of previous natural infection on vaccine response is not well known. We thus sought to evaluate the immune responses after the first and second m-RNA vaccine doses according to different groups of previous SARS-CoV-2 infection and clinical COVID-19 presentation.

## Methods

A random sample of 642 health-care workers (HCW) enrolled in a previous hospital-wide seroprevalence study of 2,590 HCW (April-2020 and November-2020; [7]) were invited to participate in this study, based on stratification of 3 groups according to their seropositive status in both surveys: 1) Naïve SARS-CoV-2 patients: Seronegative in both surveys; 2) Transient seropositivity: seropositive in first survey and negative in second survey and 3) Persistent seropositivity: seropositive in both surveys. Seropositive HCW were further classified according to the severity of the previous SARS-CoV-2 infection 1) asymptomatic; 2) moderate COVID-19, attended as out-patients; and 3) severe COVID-19: HCW that required hospital admission.

A blood sample from HCW who had received the first dose of the mRNA BNT162b2 vaccine (Pfizer/BioNTech) was obtained just before the administration of the second dose of vaccine (21 days after the first dose), and 3 weeks after the second dose.

Previous SARS-CoV-2 infection was considered when SARS-CoV-2 RT-PCR or IgG were positive (regardless of symptoms) in the first seroprevalence survey.

The SARS-CoV-2 IgG assay used is a chemiluminescent immunoanalysis of microparticles (CMIA) used for the quantitative detection of IgG antibodies against the Spike protein (RBD domain) of the SARS CoV-2 virus (IgG-S) in the Architect system (ABBOTT Diagnostics) with a cut-off <50 arbitrary Units/ml (AU/ml).

### Statistical analysis

Demographics were compared by univariate analysis using chi squared and the Student’s t test. IgG-S titers were described as geometric mean, median interquartile range. We used univariate non-parametric test, U Mann-Whitney and Krusall-Wallis test, to compare IgG-S titers between groups. Multiple comparisons were adjusted by Bonferroni method. To compare change between first and second dose we use the non parametric related sample test, Wilcoxon test.

To estimate the difference between subgroups, anti-spike IgG titer was log (natural) transformed and mixed linear regression models were performed (Supplemental Table 3). Data were evaluated with the Statistical Package SPSS-17 (IBM Armonk, NewYork) and R 3.4.4 software (https://cran.r-project.org/bin/windows/base/old/3.4.4/.)

The protocol was approved by the independent review board of the hospital.

## Results

A total of 642 HCW were invited to participate in the study, 641 accepted (99.8%) and follow-up was completed in 623 HCW (97.0%). Mean age was 45.8 (SD 10.7) years and 77.3% were women. There were not significant differences among the groups except for a lower percentage of tobacco use (14.8 vs 21.5%, p=0.029) and higher percentage of obesity (11.6 vs 6.2%, p=0.014) in patients who developed COVID-19 (Supplementary Table 1).

Median IgG-S titers were about 20-fold higher after the first dose of the vaccine in HCW with previous SARS-COV-2 antibodies (median 22,267.3 AU/ml, IQR 3,972.5-3,7091.2) than in naïve individuals (median 1,022.3 AU/ml, IQR 545.7-1,760.0, p<0.001) and even higher than those achieved after the second dose in naïve cases (median 12,873.9 AU/ml, IQR 7,472.6-19,860.2, p<.001) figure 1A, Supplementary Table 2. The boost dose in naïve subjects induced about a 12,6-fold increase in median IgG-S titers, as compared to a modest 1,27-fold increase in non-naïve cases.

**Figure 1 legend.**
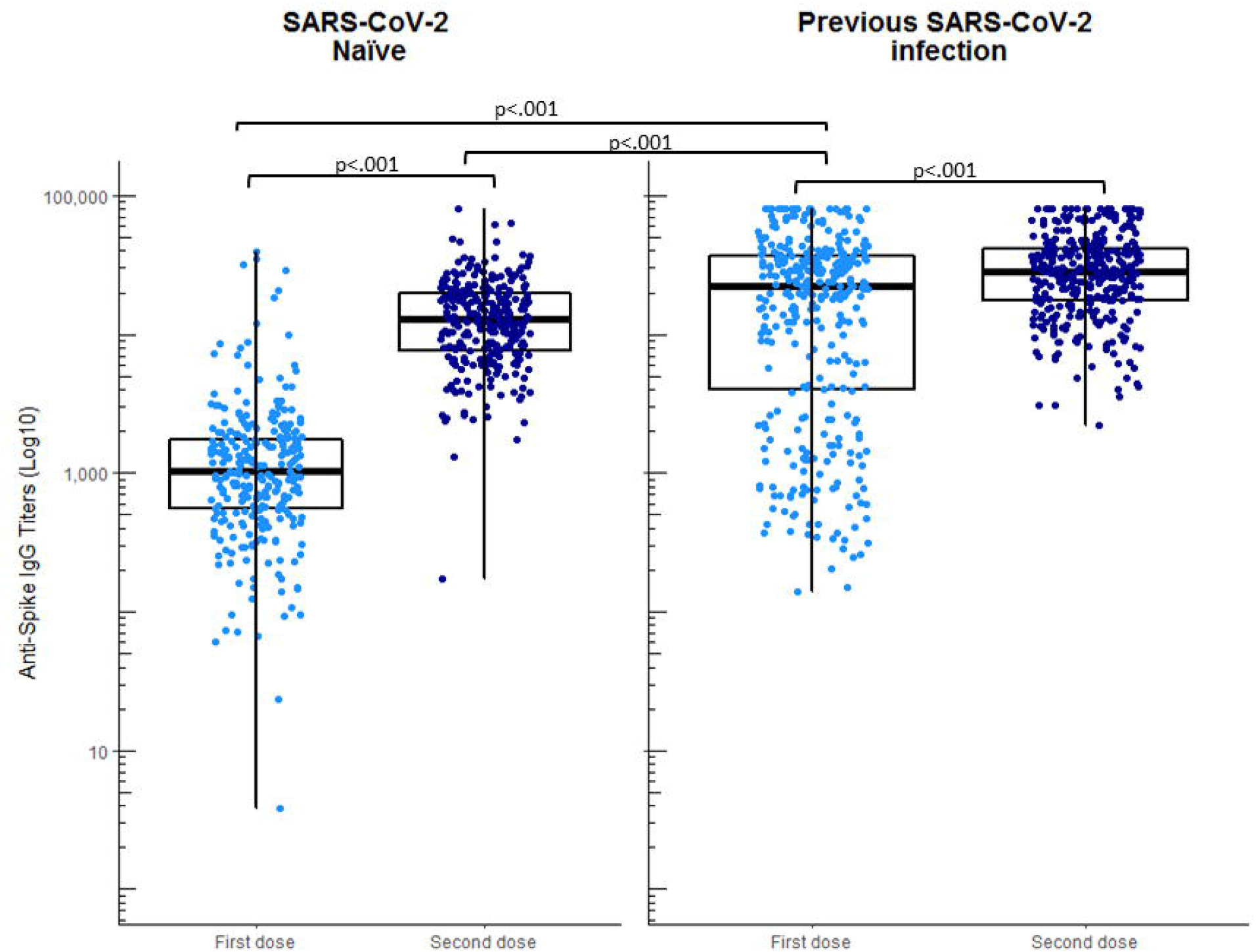
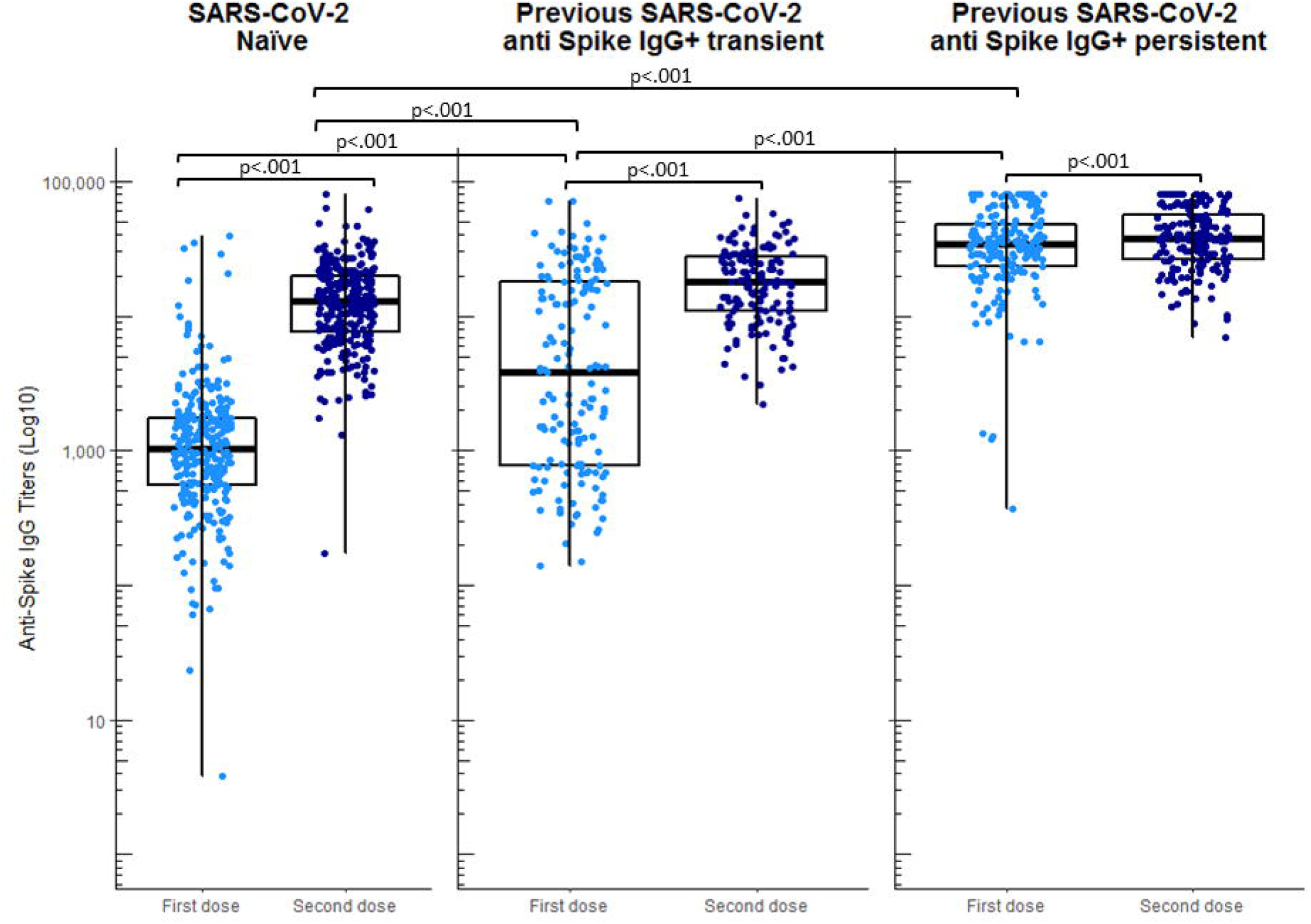
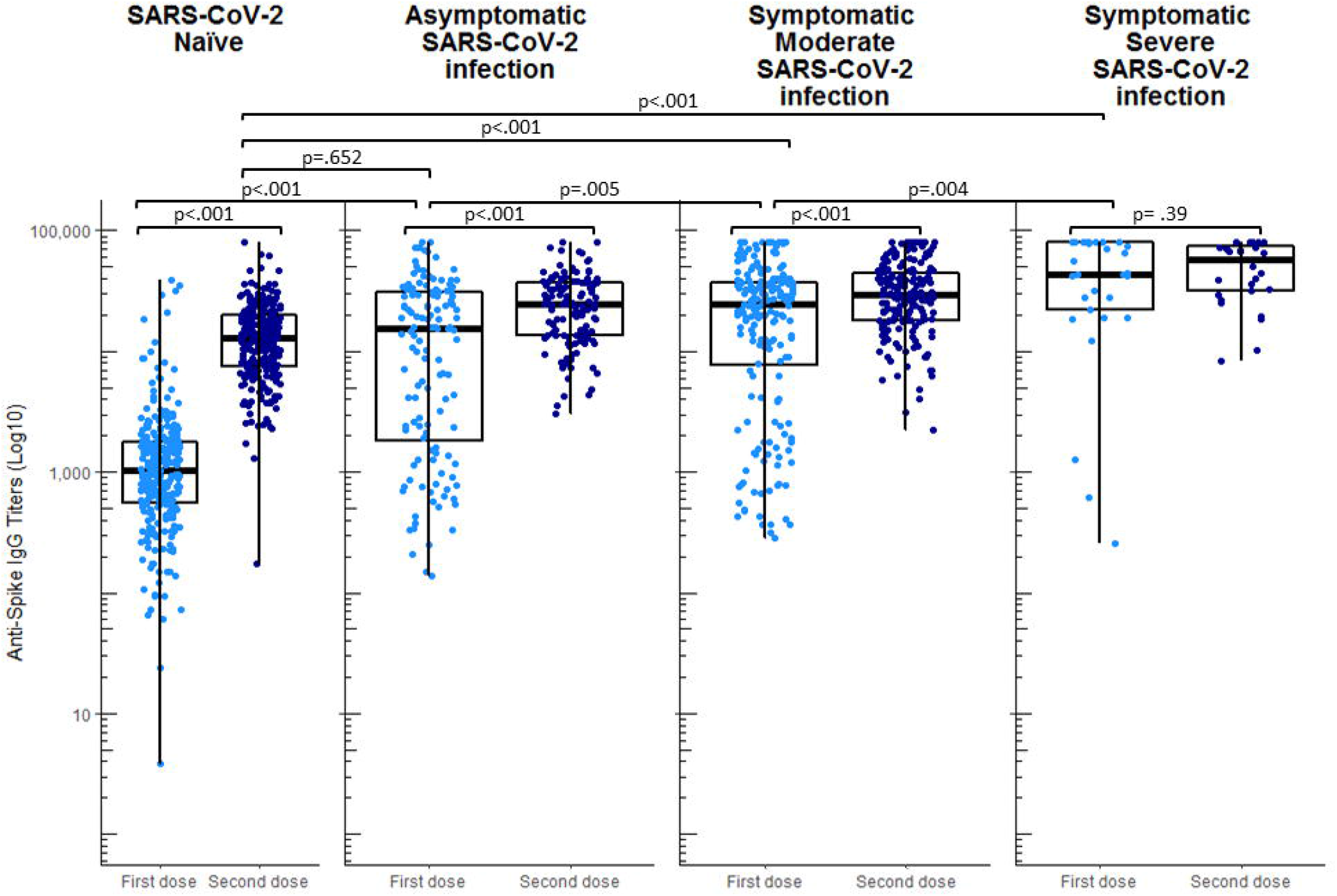
Serological response after one or two doses of the BNT162b2 mRNA COVID-19 vaccine in 641 Health Care Workers. The SARS-CoV-2 IgG anti Spike (IgG-S) were determined 3 weeks after the first (light blue dots; n=641) and 3 weeks after the second dose (dark blue dots; n=623) with a chemiluminescent immunoanalysis (Architect system, ABBOTT Diagnostics) with a cut-off <50 arbitrary Units/ml (AU/ml). IgG-S titers are represented in a logarithmic scale (Y axis). Individual data (dots), median values and interquartile range (boxes) of different groups (X axis) are depicted. Statistics: univariate non-parametric test, U Mann-Whitney and Kruskall-Wallis test, were used to compare IgG-S titers between group, as appropriate. Multiple comparisons were adjusted by the Bonferroni method. Boost effects difference for groups were evaluated by log transformed mixed linear regression models (Supplemental Table 3). **Panel A**. IgG-S titers in individuals SARS-CoV-2 naïve or with previously documented infection. Of note, the strong serologic response in previously infected individuals after the first dose was even higher than that elicited by the 2-dose vaccine protocol in naïve subjects (p<.001 for all comparisons). **Panel B**. IgG-S titers in individuals according to their serologic SARS-CoV-2 status before vaccination. Seronegative: seronegative HCW in both surveys. Transient seropositivity HCW: seropositive HCW in first survey and negative in second survey; Persistent seropositivity: seropositive HCW in both surveys. Of note, the strong serologic response in infected individuals with persistent seropositivity after the first dose was even higher than that elicited by the 2-dose vaccine protocol in naïve and transient seropositive patients (p<.001 for all comparisons); the response to the first dose of the vaccine in transient seropositive subjects was intermediate between seronegative subjects and persistent seropositive subjects. Boost effect was inversely related to the strength of the response to the first dose (p<.001). **Panel C**. IgG-S titers in individuals SARS-CoV-2 infected according to their clinical COVID-19 presentation. Of note, the strong serologic response in subjects with previous SARS-CoV-2 infection after the first dose was even higher than that elicited by the 2-dose vaccine protocol in SARS-CoV-2 naïve patients and increased progressively with the severity of the disease (p<.001). Boost effect was inversely related to the strength of the response to the first dose (p<.001)

Naïve SARS-CoV-2 individuals, transient seropositive and persistent seropositive subjects exhibited progressively higher IgG-S titers after the first dose of the vaccine (median 1,022.3 AU, IQR 545.7–1,760; median 3,860.0 AU ; IQR 781.8-18,088 and median 34,551.6 AU, IQR 23,432-48,597 respectively; p<.001 for all comparisons) figure 1B). In other words, as compared with naïve SARS-CoV-2 subjects, transient seropositive patients showed about a 3.8-fold, and persistent seropositive patients about a 33.8-fold higher level of IgG-S after the first dose of the vaccine. Interestingly, the serologic response after the first dose in transient seropositive cases was lower than that obtained with the 2-dose in naïve subjects. The boost dose induced about a 4.6-fold increase in median IgG-S titers in transient seropositive cases but a mere 1.1-fold increase in persistent seropositive subjects.

Previous COVID-19 clinical severity was also associated with increased titers of IgG-S 3 weeks after the first dose of the vaccine: asymptomatic (median 15,386.4 AU/ml, IQR 1,661.8-31,260.9), mild-moderate (median 24,388.8 AU/ml, IQR 7,158.3-37,355.5) and severe (median 43,671.7 AU/ml ; IQR 20,463.4-79,923.7) figure 1C. For all groups, median IgG-S titers in non-naïve cases after the first dose of the vaccine were higher than those achieved in naïve cases after the full 2-dose vaccination. The boost effect of the second dose was lower for symptomatic cases as compared to asymptomatic subjects (Supplementary Table 2).

A total of 13 subjects developed symptomatic COVID-19 after the first dose of the vaccine, showing a median IgG-S titer of 2,485.8 AU/ml, (IQR 1,144.6-9,842.9). Since it is impossible to discern the relative contribution of the vaccine and post-vaccine SARS-CoV-2 infection to the IgG titer, no comparison was made between this group and others.

We further evaluated the different serological response in the groups mentioned above with a linear regression model after a logarithm transformation of IgG titers and adjusting for age, sex and comorbidity. Qualitatively the results are essentially unchanged. Previous SARS-CoV-2 exposure and disease severity was associated with strong serologic response after the first dose of at least a similar magnitude as the full 2-dose vaccine in SARS-CoV-2 naïve subjects (Supplementary Table 3). Boosting dose effect was lower for groups with a strong response to the first dose.

## Discussion

Our serological evaluation in HCW shows a dramatic differential response 3-weeks after a single dose of the BNT162b2 mRNA vaccine according to previous SARS-CoV-2 infection and serological status. So far, this is the large study on immune response after first and second dose of BNT162b2 vaccine. Of note patients with previous history of SARS-CoV-2 infection reached at least one order of magnitude higher titers of anti Spike-IgG. Interestingly, IgG-S titers achieved by a single dose in seropositive patients was higher than those achieved by a full vaccination in SARS-CoV-2 naïve cases.

Although we have not assessed the presence of neutralizing antibodies, our results reinforce preliminary reports with a small number of cases [3–6]. This robust serologic response is highly correlated to the presence of neutralizing antibodies [8]. However, at present, we have no data regarding the duration of the serologic response nor the degree of protection against SARS-CoV-2 infection.

Our work provides some additional interesting data. First HCW who had lost their SARS-CoV-2 antibodies exhibit a lower response, that might be insufficient to provide SARS-CoV-2 protection[3,8]. Conversely, the combination of clinical COVID-19 and the presence of anti-Spike-IgG SARS-COV-2 antibodies before vaccination are strong predictors of a robust serologic response to a single dose of the mRNA vaccine. This response is of similar magnitude or higher than the one achieved by a 2-dose vaccination in naïve SARS-CoV-2 subjects. Our data suggest that the serologic response is close to saturation in seropositive patients after a single mRNA vaccine dose, as suggested by a very limited boost response to a second dose of the vaccine.

There is current controversy about the most efficient strategy regarding the use of available vaccines in order to offer a wider population protection[9,10]. In this regards results from the UK suggest a decrease of COVID-19 following a program of single dose vaccination [11]. In addition, this strategy may reduce the exacerbated clinical symptoms associated with the second dose in seropositive individuals [5]. Whether enhanced single dose mRNA vaccine-induced serologic response among previously seropositive individuals will show a lasting response as compared to boosted vaccines is currently unknown. As no trial has evaluated the medium-term protective effects of a single-dose mRNA protocol, health policy makers rely on surrogate markers such as SARS-CoV-2 seropositivity prior to vaccination, for this difficult decision [12]. Our results suggest that deferring the second dose of the vaccine to seropositive individuals with a predictable strong serologic response may not a high risk strategy for them, offering the option for vaccination of other individuals at risk until a wider vaccine availability allows for a standard 3-4 week interval between mRNA vaccine doses as this is the protocol that has been tested in clinical trials. If SARS-CoV-2 serology is available, seropositive patients may be offered a deferred boost vaccination. In circumstances where serology is unknown, previous COVID-19 severity may guide the selection of patients.

## Data Availability

Full data are available at our hospital upon request

## Acknowledgments

We thank all workers of the Hospital Universitario Fundación Alcorcón for their everyday work and cooperation in this study.

## References

1. World Health Organization. Background document on the mRNA vaccine BNT162b2 (Pfizer-BioNTech) against COVID-19: background document to the WHO interim recommendations for use of the Pfizer–BioNTech COVID-19 vaccine, BNT162b2, under emergency use listing, 14 January 2021. 2021; Available at: https://apps.who.int/iris/handle/10665/338671. Accessed 8 March 2021.

2. Polack FP, Thomas SJ, Kitchin N, et al. Safety and Efficacy of the BNT162b2 mRNA Covid-19 Vaccine. N Engl J Med 2020; Ahead of print DOI: 10.1056/NEJMoa2034577

3. Manisty C, Otter AD, Treibel TA, et al. Antibody response to first BNT162b2 dose in previously SARS-CoV-2-infected individuals. The Lancet 2021; Ahead of print DOI: 10.1016/S0140-6736(21)00501-8

4. Prendecki M, Clarke C, Brown J, et al. Effect of previous SARS-CoV-2 infection on humoral and T-cell responses to single-dose BNT162b2 vaccine. The Lancet 2021; Ahead of print DOI:10.1016/S0140-6736(21)00502-X

5. Krammer F, Srivastava K, Alshammary H, et al. Antibody Responses in Seropositive Persons after a Single Dose of SARS-CoV-2 mRNA Vaccine. N Engl J Med 2021; Ahead of print DOI:10.1056/NEJMc2101667.

6. Saadat S, Tehrani ZR, Logue J, et al. Binding and Neutralization Antibody Titers After a Single Vaccine Dose in Health Care Workers Previously Infected With SARS-CoV-2. JAMA 2021; Ahead of print DOI: 10.1001/jama.2021.3341.

7. Galán MI, Velasco M, Casas ML, et al. Hospital-Wide SARS-CoV-2 seroprevalence in health care workers in a Spanish teaching hospital. Enferm Infecc Microbiol Clin 2020; Ahead of print DOI: 10.1016/j.eimc.2020.11.015

8. Bartsch YC, Fischinger S, Siddiqui SM, et al. Discrete SARS-CoV-2 antibody titers track with functional humoral stability. Nat Commun 2021; 12:1018.

9. Bieniasz P. The case against delaying SARS-CoV-2 mRNA vaccine boosting doses. Clin Infect Dis 2021; Ahead of print Doi: 2021. 10.1093/cid/ciab070

10. Kadire SR, Wachter RM, Lurie N. Delayed Second Dose versus Standard Regimen for Covid-19 Vaccination. N Engl J Med 2021; 384:e28. Ahead of print DOI: 10.1056/NEJMclde2101987.

11. Vasileiou E, Simpson CR, Robertson C, et al. Effectiveness of First Dose of COVID-19 Vaccines Against Hospital Admissions in Scotland: National Prospective Cohort Study of 5.4 Million People. SSRN Electron J 2021; Ahead of print DOI: 10.2139/ssrn.3789264

12. Bubar KM, Reinholt K, Kissler SM, et al. Model-informed COVID-19 vaccine prioritization strategies by age and serostatus. Science 2021; Ahead of print DOI: 10.1126/science.abe6959

