## Supplemental tables for "Impact of previous COVID-19 on immune response after a single dose of BNT162b2 SARS-CoV-2 vaccine"

**Supplementary tables**

**Supplementary Table 1. Participant characteristics**

|  | **Total** | **No COVID-19** | **COVID-19 positive** | **Univariate test** |
| --- | --- | --- | --- | --- |
| **VARIABLE** | **no=641** | **no=284** | **no=357** |  |
| **Women** | 495 (77.2%) | 227 (79.9%) | 268 (75.1%) | 0.145 |
| **Age (mean ± SD)* years** | 45.5 ± 10.6 | 45.4 ± 10.4 | 45.6 ± 10.9 | 0.747 |
| **Clinical conditions** |  |  |  |  |
| Tobacco use | 114 (17.8%) | 61 (21.5%) | 53 (14.8%) | **0.029** |
| Chronic lung disease | 74 (11.5%) | 37 (13%) | 37 (10.4%) | 0.294 |
| High blood pressure | 44 (6.9%) | 21 (7.4%) | 23 (6.4%) | 0.636 |
| Obesity | 55 (8.6%) | 33 (11.6%) | 22 (6.2%) | **0.014** |
| Other cardiovascular disease | 25 (3.9%) | 8 (2.8%) | 17 (4.8%) | 0.206 |
| Diabetes mellitus | 18 (2.8%) | 7 (2.5%) | 11 (3.1%) | 0.639 |
| Immunodeficiency | 10 (1.6%) | 6 (2.1%) | 5 (1.4%) | 0.757 |
| Cancer | 5 (0.8%) | 4 (1.4%) | 1 (0.3%) | 0.176 |
| Liver disease | 2 (0.3%) |  | 2 (0.6%) | 0.506 |
| Chronic kidney disease | 1 (0.2%) | 1 (0.4%) |  | 0.443 |
| **Previous SARS-CoV-2 infection** |  |  |  |  |
| Total | 357 |  | 357 |  |
| Asymptomatic | 132 |  | 132 |  |
| Symptomatic Moderate COVID | 196 |  | 196 |  |
| Symptomatic Severe COVID | 29 |  | 29 |  |

*SD: standard deviation

COVID-19: Health care workers with previous SARS-CoV-2 documented infection

**Supplementary Table 2. SARS-CoV-2 titer by serologic and symptomatic group**

| **SUBGROUP** |  | **Descriptive statistics** | | | | **First doce** | | | | | **Univariate paired samples test (dose1-dose2)** |
| --- | --- | --- | --- | --- | --- | --- | --- | --- | --- | --- | --- |
|  | **Dose** | **N** | **Geometric mean** | **Median** | **Interquartile range** | **Univariate test** | **Univariate post hoc test adjusted by Bonferroni Method** | | **Univariate post hoc test adjusted by Bonferroni Method** | |  |
|  |  |  |  |  |  |  | **vs Naïve** | **vs transient** | **vs asymptomatic** | **vs symptomatic moderate** |  |
| **SARS-CoV-2** |  |  |  |  |  |  |  |  |  |  |  |
| Naïve | First | 284 | 992.4 | 1022.3 | 545.7 - 1760 | <0.001* |  |  |  |  | <0.001*** |
|  | Second | 279 | 11847.8 | 12873.9 | 7472.6 - 19860.2 |  |  |  |  |  |  |
| COVID-19 positive (PCR/IgG/Ag) | First | 357 | 11721.3 | 22267.3 | 3972.5 - 37091.2 |  |  |  |  |  | <0.001*** |
|  | Second | 344 | 26079.6 | 28355.1 | 17569.1 - 41120.8 |  |  |  |  |  |  |
| **Previous SARS-CoV-2 serology** |  |  |  |  |  |  |  |  |  |  |  |
| anti Spike IgG+ transient | First | 150 | 3664.8 | 3860.0 | 781.8 - 18088.1 | <0.001* | <0.001* |  |  |  | <0.001*** |
|  | Second | 146 | 16886.0 | 17919.4 | 10931.4 - 27918.2 |  |  |  |  |  |  |
| anti Spike IgG+ persistent | First | 194 | 31526.0 | 34551.6 | 23432 - 48597 |  | <0.001* | <0.001* |  |  | <0.001*** |
|  | Second | 188 | 37122.0 | 37814.2 | 26291.1 - 56357.7 |  |  |  |  |  |  |
| **Previous SARS-CoV-2 symptoms** |  |  |  |  |  |  |  |  |  |  |  |
| Asymptomatic | First | 132 | 7763.2 | 15386.4 | 1661.8 - 31260.9 | <0.001** | <0.001* |  |  |  | <0.001*** |
|  | Second | 129 | 21686.5 | 24458.4 | 13361.6 - 37067.2 |  |  |  |  |  |  |
| Symptomatic Moderate | First | 196 | 13515.7 | 24388.8 | 7158.3 - 37355.5 |  | <0.001* |  | 0.005* |  | <0.001*** |
|  | Second | 187 | 27364.9 | 29171.6 | 18101.3 - 45229.3 |  |  |  |  |  |  |
| Symptomatic Severe | First | 29 | 29198.7 | 43671.7 | 20463.4 - 79923.7 |  | <0.001* |  | <0.001* | 0.004* | 0.39*** |
|  | Second | 28 | 44245.0 | 57910.0 | 30145.8 - 78282.3 |  |  |  |  |  |  |
| *Univariate U Mann Whitney test; **Univariate Kruskall Wallis test; ***Univariate Wilcoxon test | | | | | | | | | | | |

Previous SARS-CoV-2 anti Spike IgG+ Transient seropositivity: seropositive in first survey and negative in second survey

Previous SARS-CoV-2 anti Spike IgG+ Persistent seropositivity: seropositive in both surveys

**Supplementary table 3A.**

| **MODEL** | **Subgroup** | **Unadjusted estimation** | | | | | | | | **Estimation adjusted by age, gender and comorbidity** | | | | | | |
| --- | --- | --- | --- | --- | --- | --- | --- | --- | --- | --- | --- | --- | --- | --- | --- | --- |
|  |  | **Coefficient** | **CI95%** | | | **p-value** | **Relative difference** | **CI95%** | | **Coefficient** | **CI95%** | | **p-value** | **Relative difference** | **CI95%** | |
| **Model 1** | **COVID** |  |  | |  |  |  |  |  |  |  |  |  |  |  |  |
|  | Naïve | reference |  | |  |  |  |  |  |  |  |  |  |  |  |  |
|  | Previous infection | 2.5 | 2.2 | | 2.7 | <0.001 | 11.8 | 9.4 | 14.7 | 2.4 | 2.2 | 2.7 | <0.001 | 11.4 | 8.8 | 14.8 |
| **Model 2** | **Previous SARS-CoV-2 serology** |  |  | |  |  |  |  |  |  |  |  |  |  |  |  |
|  | Naïve | reference |  | |  |  |  |  |  |  |  |  |  |  |  |  |
|  | anti Spike IgG+ transient | 1.3 | 1.1 | | 1.5 | <0.001 | 3.6 | 2.9 | 4.6 | 1.3 | 1.1 | 1.6 | <0.001 | 3.8 | 2.9 | 4.9 |
|  | anti Spike IgG+ persistent | 3.5 | 3.2 | | 3.7 | <0.001 | 31.8 | 25.6 | 39.4 | 3.4 | 3.2 | 3.7 | <0.001 | 31.2 | 24.4 | 39.9 |
| **Model 3** | **Previous SARS-CoV-2 symptoms** |  |  | |  |  |  |  |  |  |  |  |  |  |  |  |
|  | Asymptomatic | reference |  | |  |  |  |  |  |  |  |  |  |  |  |  |
|  | Symptomatic Moderate | .5 | .2 | | .9 | .002 | 1.7 | 1.2 | 2.5 | .7 | .4 | 1.1 | <0.001 | 2.1 | 1.4 | 3.1 |
|  | Symptomatic Severe | 1.3 | .7 | | 2.0 | <0.001 | 3.8 | 2.0 | 7.1 | 1.9 | 1.1 | 2.6 | <0.001 | 6.5 | 3.2 | 13.3 |
|  | Log transformed linear regression models | | |  |  |  |  |  |  |  |  |  |  |  |  |  |
|  | Relative difference: exponentiated regression coefficients | | | | |  |  |  |  |  |  |  |  |  |  |  |

**Supplementary Table 3B.**

| **MODEL** | **Subgroup** | **Interaction time*subgroup effect** | | | | **Model estimated means** | | | | | | **Time pairwise comparison (Bonferroni adjustment for multiple comparisons)** | | | | |
| --- | --- | --- | --- | --- | --- | --- | --- | --- | --- | --- | --- | --- | --- | --- | --- | --- |
|  |  | **p-value** | **Estimated paremeter** | **CI95%** | | **Dose 1** | **CI95%** | | **Dose 2** | **CI95%** | | **Differenci dose2-dose1** | **CI95%** | | **p-value** | **Relative difference** |
| **Model 1** | **COVID** | <0.001 |  |  |  |  |  |  |  |  |  |  |  |  |  |  |
|  | Naïve |  | ref |  |  | 6.91 | 6.71 | 7.11 | 9.39 | 9.28 | 9.49 | 2.48 | 2.34 | 2.62 | <0.001 | 11.94 |
|  | Previous infection |  | 1.68 | 1.49 | 1.86 | 9.34 | 9.17 | 9.51 | 10.14 | 10.05 | 10.23 | 0.80 | 0.68 | 0.92 | <0.001 | 2.23 |
| **Model 2** | **Previous SARS-CoV-2 serology** | <0.001 |  |  |  |  |  |  |  |  |  |  |  |  |  |  |
|  | Naïve |  | ref |  |  | 6.92 | 6.75 | 7.08 | 9.40 | 9.30 | 9.49 | 2.48 | 2.36 | 2.60 | <0.001 | 11.94 |
|  | anti Spike IgG+ transient |  | 0.98 | 0.78 | 1.18 | 8.25 | 8.04 | 8.46 | 9.75 | 9.63 | 9.87 | 1.50 | 1.35 | 1.66 | <0.001 | 4.49 |
|  | anti Spike IgG+ persistent |  | 2.33 | 2.14 | 2.52 | 10.35 | 10.16 | 10.54 | 10.50 | 10.39 | 10.61 | 0.15 | 0.01 | 0.29 | 0.037 | 1.16 |
| **Model 3** | **Previous SARS-CoV-2 symptoms** | <0.001 |  |  |  |  |  |  |  |  |  |  |  |  |  |  |
|  | Asymptomatic |  | ref |  |  | 8.78 | 8.48 | 9.08 | 9.91 | 9.77 | 10.04 | 1.13 | 0.91 | 1.35 | <0.001 | 3.10 |
|  | Symptomatic Moderate |  | 0.46 | 0.18 | 0.74 | 9.52 | 9.27 | 9.77 | 10.19 | 10.07 | 10.30 | 0.67 | 0.49 | 0.85 | <0.001 | 1.95 |
|  | Symptomatic Severe |  | 0.94 | 0.42 | 1.47 | 10.55 | 9.90 | 11.20 | 10.74 | 10.45 | 11.02 | 0.19 | -0.29 | 0.67 | 0.44 | 1.21 |
|  | Multivariate mixed model: including sex, age, comorbidity, time, subgroup and time*subgroup interaction | | | | | | | | | | | | | | | |
|  | Natural log transformed mixed linear regression models | | | | | | | | | | | | | | | |
|  | Relative difference: exponentiated regression coefficients | | | | | | | | |  |  |  |  |  |  |  |

To estimate the difference between subgroups, anti-spike IgG titer was log (natural) transformed and mixed linear regression models were performed. These models include time as repeated measure, group as fixed effect and the interaction term time*group. Sex, gender and comorbidity were included to adjust the estimated effect. An interaction term statistically significant was interpreted as difference between first and second dosis depend of group. The arithmetic mean of log-transformed values corresponds to the log geometric mean of the original values. So, the exponentiated regression coefficient estimated in model can be interpreted as relative change in the geometric mean of anti-spike IgG titer in original scale.
